# Protocol for Systematic Review and Meta-Analysis of Prehospital Large Vessel Occlusion Screening Scales

**DOI:** 10.1101/2024.08.19.24312257

**Authors:** Hidetaka Suzuki, Yohei Okada, Yamagami Hiroshi, Mikito Hayakawa, Yoshimoto Takeshi, Hitoshi Kobata

## Abstract

**Background:** Large Vessel Occlusion (LVO) is a serious condition that causes approximately 24-46% of acute ischemic strokes (AIS). LVO strokes tend to have higher mortality rates and result in more severe longterm disabilities compared to non LVO ischemic strokes. Early intervention with endovascular therapy (EVT) is recommended; however, EVT is limited to tertiary care hospitals with specialized facilities. Therefore, identifying patients with a high probability of LVO in prehospital settings and ensuring their rapid transfer to appropriate hospitals is crucial. While LVO diagnosis typically requires advanced imaging like MRI or CT scans, various scoring systems based on neurological symptoms have been developed for prehospital use. Although previous systematic reviews have addressed some of these scales, recent studies have introduced new scales and additional data on their accuracy. This systematic review and meta-analysis aim to summarize the current evidence on the diagnostic accuracy of these prehospital LVO screening scales.

**Methods:** This systematic review and meta-analysis will be conducted in accordance with the PRISMA-DTA Statement and the Cochrane Handbook for Systematic Reviews of Diagnostic Test Accuracy. We will include observational studies and randomized controlled trials that assess the utility of LVO scales in suspected stroke patients in prehospital settings. Eligible studies must provide sufficient data to calculate sensitivity and specificity, and those lacking such data or being case reports will be excluded. The literature search will cover CENTRAL, MEDLINE, and Ichushi databases, including studies in English and Japanese. Bias will be assessed using QUADAS-2, and meta-analysis will be conducted using a random effects model, with subgroup and sensitivity analyses to explore heterogeneity.

## Background

Large Vessel Occlusion (LVO) is a severe condition characterized by the occlusion of major cerebral arteries, accounting for 24–46% of acute ischemic strokes (AIS). Compared to non-LVO ischemic strokes, LVO strokes are associated with higher mortality rates and a greater risk of long-term disability [1, 2]. Endovascular therapy (EVT) plays a crucial role in the treatment of LVO, with guidelines recommending early intervention[3]. However, capability of EVT is limited to the tertiary care hospitals with specialized team and facility; thus, it is essential to identify the candidates with a high probability of LVO in the prehospital setting and ensure their rapid transfer to appropriate hospitals.

Generally, the definitive diagnosis of LVO relies on advanced imaging techniques such as MRI or CT scans [1]. However, to recognize the patients with LVO in the prehospital setting, there are various scoring systems using neurological symptoms and the degree of consciousness. Previously, some systematic reviews on the scores were published; however, since then several papers reported the novel scores and further information about their accuracy and validity. Currently, there is few latest summarized evidence on the diagnostic accuracy of the scoring system. This systematic review and meta-analysis aim to summarize the evidence of these prehospital screening scales currently available or already in use.

## Methods

### Disclosure of Study Design and Protocol

We plan to conduct a systematic review and meta-analysis of diagnostic accuracy, adhering to the PRISMA-DTA Statement and the Cochrane Handbook for Systematic Reviews of Diagnostic Test Accuracy. The review protocol will be registered on the UMIN clinical trial registry and posted on a preprint server (medRxiv).

### Criteria for Study Inclusion

1. **Study Design**: Retrospective or prospective observational studies and randomized controlled trials assessing the utility of LVO scales in suspected stroke patients in prehospital settings. We will include the studies providing sufficient detail to calculate 2×2 table, specificity and sensitivity. The studies lacking data for 2×2 tables or those that are case reports or case-control studies (two-gate study) will be excluded.
2. **Participants**: The study participant will be patients suspected of having a stroke in the prehospital phase, with no age restrictions.
3. **Setting**: The eligible study setting is involving prehospital settings such as emergency medical services (EMS) or physician-staffed ambulance/helicopter.
4. **Index Tests**: We will include all various LVO scales usable in prehospital settings, such as ELVO screen, RACE, CPSSS, LAMS, 3-item stroke scale, FACE2AD, JUST, and JUST7, with all cutoff values included.
5. **Target Condition**: The target condition is an ischemic stroke due to LVO (occlusion of the internal carotid artery, middle cerebral artery M1 or M2, or basilar artery).
6. **Reference Standard**: We will set the reference standard as the results from following modality CTA(Computed tomography angiography), MRA(magnetic resonance angiography), invasive angiography, TCD(Transcranial doppler), or the original standard reference employed in each study.

### Literature Search and Selection

Since a systematic review regarding the LVO scales was published previously, this review will be accordance with the concept of updating the review. The eligible literature reported before the previous systematic review was published will be identified from the previous systematic review. Eligible literature published after the previous systematic review will be identified by searching the medical database as below. The search will cover studies published in English and Japanese, including abstracts from conferences. We will search the following databases CENTRAL, MEDLINE, and Ichushi. The search strategy will be derived from the strategy used in the previous literature. Additional references will be identified by hand-search, and we will attempt to the contributing authors to obtain the additional information if necessary.

### Data Collection

Following data will be extracted independently by two review authors: study design, participants, index tests, target conditions, outcomes, diagnostic accuracy, authors, country, setting, funding and conflict of interest statement. Discrepancies will be resolved through discussion or consultation with a third author.

### Bias Assessment

Two authors will independently assess the methodological quality using QUADAS-2, evaluating the risk of bias in patient selection, index tests, reference standards, and flow and timing.

### Meta-Analysis

True positives, false positives, true negatives, and false negatives will be extracted from each study. Analyses will be stratified by cutoff values of each LVO scale. Heterogeneity among studies will be assessed using the I^2^ statistic, with a random effects model accounting for bias.

### Subgroup Analyses

Analyses will consider differences in evaluator of index test, study design, reference standards, age, and subtype or symptom of the stroke patients.

### Sensitivity Analyses

To examine the impact of major methodological factors or decisions on the primary results, sensitivity analyses will only include the studies with low risk of bias and differences in study year.

## Data Availability

All data produced in the present study are available upon reasonable request to the authors.

